# Uncertainty-Aware Multiscale Treatment Response Forecasting in Advanced Non-Small Cell Lung Cancer Using Residual-Variance Conformal Prediction

**DOI:** 10.64898/2026.09.30.26364431

**Authors:** Faisal Yaseen, Daniel S. Hippe, Sunan Cui, Jie Fu, Shouyi Wang, Lei Deng, Ting Ye, Paul E. Kinahan, Jing Zeng, John H. Gennari, Stephen R. Bowen

**Affiliations:** Department of Biomedical Informatics and Medical Education, University of Washington, Seattle, WA, USA; Department of Radiation Oncology, University of Washington, Seattle, WA, USA; Fred Hutchinson Cancer Center, Seattle, WA, USA; Department of Biostatistics, University of Washington, Seattle, WA, USA; Department of Radiology, University of Washington, Seattle, WA, US; The University of Texas at Arlington, Arlington, TX, USA

**Keywords:** FDG-PET/CT, non-small cell lung cancer, voxel-level prediction, conformal prediction, uncertainty quantification, prediction intervals, spatial statistics, variogram

## Abstract

Voxel-level prediction of treatment response from longitudinal Fluorodexoyglucose (FDG) positron emission tomography / computed tomography (PET/CT) can enable spatially adaptive dose planning in advanced non-small cell lung cancer (NSCLC), but point predictions alone lack the uncertainty quantification needed for safe clinical decision-making. Standard conformal prediction (CP) provides distribution-free coverage but yields spatially uniform intervals and assumes exchangeability—an assumption violated by intra-tumoral spatial correlation. We propose a multiscale CP framework built on a re-optimized VoxelForecast generalized least squares model that explicitly captures voxel-level covariance. Variogram sensitivity analysis identified the Stable model as significantly superior to previouly used Matérn family. The framework combines hierarchical nested leave-one-patient-out calibration, which restores exchangeability for valid voxel-level coverage, with residual-variance CP (VarCP), which learns a heteroscedastic, location-dependent scale to produce adaptive intervals while preserving finite-sample marginal coverage guarantees. On two prospective NSCLC trial cohorts—FLARE-RT (N=25 locally advanced, chemoradiotherapy; 11,100 voxels) and BRIGHT (N=19 metastatic, chemoimmunotherapy; 26,980 voxels)—multiscale feature integration reduced voxel-level prediction error by 35% and 31%, respectively. VarCP maintained nominal coverage while producing 7–18% (FLARE-RT) and 4–8% (BRIGHT) narrower voxel-level intervals than a hierarchically calibrated standard CP baseline (paired Wilcoxon signed-rank, significant at most miscoverage levels), with comparable lesion-level gains (7–18% and 6–10%) via median-based aggregation of voxel residuals. VarCP delivers tighter yet mathematically guaranteed spatial uncertainty maps, supporting both voxel-level dose-adjustment and whole-lesion treatment planning in advanced NSCLC.

## I Introduction

Non-Small Cell Lung Cancer (NSCLC) remains a leading cause of cancer-related mortality, requiring treatment strategies tailored to disease stage [1]. For patients with locally advanced NSCLC (laNSCLC), in which disease is confined to the thorax without distant metastases, the standard of care is concurrent chemoradiotherapy followed by immunotherapy in eligible patients. In contrast, first-line-treatment for metastatic NSCLC (mNSCLC), characterized by distant metastatic spread, relies heavily on systemic chemo-immunotherapy (chemoICI) [1]. Furthermore, in mNSCLC patients presenting with oligoprogressive disease, addition of localized radiotherapy is commonly used to prolong progression-free survival and overall survival [2]–[4]. Despite these established therapeutic pathways, clinical benefit is limited and subject to toxicity. One important driver of this variability is the profound spatial heterogeneity of the disease, both within individual lesions (intra-lesional) and across different lesions within the same patient (inter-lesional). Distinct subregions within a primary tumor and different metastatic lesions in the same patient often display highly discordant, spatially localized responses to otherwise uniform systemic and local therapies [5], [6]. Standard treatment response assessment techniques, which rely on imaging-derived size and intensity metrics summarized over the entire tumor, often overlook this underlying spatial heterogeneity, resulting in imprecise response characterization that may limit optimal treatment decision-making [7], [8]. Therefore, accurately predicting treatment response across multiple spatial and temporal scales remains a critical unmet clinical need.

Longitudinal fluorodeoxyglucose–positron emission tomography/computed tomography (FDG PET/CT) has been widely used to characterize spatial variability in tumor biology, enabling noninvasive quantification of intra- and inter-lesional metabolic heterogeneity over time, as measured by the standardized uptake value (SUV) [9]. Prior work by *Aerts et al*. [10] using FDG PET/CT demonstrated that intra-tumor heterogeneity, characterized by spatial variation in metabolic activity across tumor regions, is a strong prognostic indicator of patient outcomes. In their framework, heterogeneity was quantified using whole tumor radiomic features that summarize voxel intensity distributions and texture patterns into lesion-level descriptors. While these macroscopic features capture global tumor phenotype, their reliance on spatial aggregation averages local variation across the tumor volume, limiting the ability to preserve fine scale voxel-level heterogeneity that may represent biologically distinct subregions with differential treatment response. More granular approaches, including subvolume metabolic control probability modeling [11] and voxel-wise regression for recurrence prediction [12], moved beyond whole-tumor aggregation but still treated voxels as spatially independent and did not jointly model patient- and voxel-level predictors. In our previous work [13], we showed that combining patient- and voxel-level baseline features while accounting for voxel-level spatial correlation improves point prediction accuracy of voxel-level treatment response. That framework modeled spatial correlation using a Matérn variogram, though the choice of variogram model was not systematically evaluated to improve prediction accuracy. A systematic variogram sensitivity analysis is needed to identify a variogram that improves prediction accuracy. However, in high-stakes clinical decision-making such as spatially adaptive dose-escalation, predictive accuracy alone is insufficient. For safe integration into clinical decision-making, predictive models must be accompanied by robust uncertainty quantification that allows clinicians to gauge the reliability of the model [14].

Conformal prediction (CP) is a non-parametric framework for uncertainty quantification. Unlike Bayesian or bootstrap-based approaches, CP requires no parametric assumptions about the data-generating distribution. Instead, it provides prediction intervals with finite-sample coverage guarantees under the mild assumption of exchangeable observations, meaning the joint distribution of the observation sequence is invariant under permutations, a condition implied by *i*.*i*.*d*. sampling [15]. This assumption does not hold when modeling voxel-level treatment response, because neighboring voxels within a tumor are inherently correlated due to shared underlying biology. In voxel-level treatment response prediction, the clinical importance of uncertainty varies across the tumor volume. Voxels in spatially homogeneous regions, where neighboring tissue shares similar metabolic behavior, can be predicted with relatively low expected prediction error, while voxels at tumor boundaries, near necrotic cores, or in regions of mixed tissue composition exhibit far greater biological variability and are more likely to respond unpredictably to therapy [13], [16]. Despite this inherent spatial heterogeneity, standard CP produces uniform prediction intervals across all voxels, treating every location identically regardless of its local spatial context or clinical relevance [15], [17].

In this work, we develop a voxel-level conformal prediction framework for FDG PET/CT response that combines a hierarchical nested leave-one-out calibration strategy with a learned residual-variance model. First, we employ a nested leave-one-patient-out design that respects the hierarchical structure of voxels within patients, restoring the exchangeability required for valid voxel-level conformal calibration. Second, building on the normalized CP framework of Lei *et al*. [18] and Romano *et al*. [19], and motivated by the greater residual variability expected at tumor boundaries, we train a residual-variance model *σ*(*x*) that predicts the magnitude of log-residuals from the same multiscale features used for the predictive model, yielding heteroscedastic (location-dependent) residual-variance conformal prediction (VarCP) intervals. This yields voxel-level intervals that adapt to predicted local difficulty while preserving the standard finite-sample marginal coverage guarantees of CP. We benchmark VarCP against standard (homoscedastic) conformal prediction (StdCP), which uses a single global residual quantile and produces non-adaptive, fixed-width intervals across voxels.

We validated our proposed framework on longitudinal FDG PET/CT data from two independent prospective clinical trials representing distinct disease stages and treatment regimens. The main contributions of this paper are:

1. A systematic variogram sensitivity analysis evaluating multiple variogram models for voxel-level treatment response prediction, identifying the Stable variogram model as optimal and improving upon the Matérn model used in prior work [13].
2. A unified voxel-level prediction framework that integrates multiscale (patient- and voxel-level) baseline features within the Voxel Forecast model, demonstrating substantial reductions in prediction error compared with patient-level-only models.
3. A residual-variance conformal prediction (VarCP) method for voxel-level FDG PET/CT treatment response, which leverages a learned heteroscedastic scale model to construct adaptive prediction intervals. VarCP was empirically benchmarked against standard conformal prediction (StdCP), yielding consistently tighter voxel-level intervals with similar or slightly higher coverage.
4. A multiscale evaluation demonstrating how voxel-level prediction intervals translate to empirical lesion-level coverage through median-based aggregation, providing complementary clinical decision support at both the voxel and lesion (whole-tumor treatment) scales.

The remainder of this paper is organized as follows. Section II describes the study cohorts, data preprocessing, the Voxel Forecast spatial prediction framework with re-optimized variogram modeling, and the conformal prediction framework including both StdCP and the proposed VarCP. Section III presents the experimental results, validated across two independent prospective NSCLC trial cohorts spanning distinct disease stages and treatment paradigms (chemoradiotherapy vs. chemoimmunotherapy), including variogram sensitivity analysis, voxel-level and lesion-level coverage, and qualitative patient examples illustrating the clinical interpretation of voxel-level and lesion-level intervals. Section IV discusses the findings, clinical implications, limitations, and directions for future work.

## II METHODOLOGY

### A Study Cohorts

We utilized longitudinal imaging data from the following two distinct patient cohorts with non-small cell lung cancer (NSCLC) representing distinct disease stage and therapy regimens: (1) locally advanced NSCLC (laNSCLC) participants from the Functional Lung Avoidance and Response-adaptive Escalation Radiation Therapy trial (FLARE-RT, NCT02773238), and (2) metastatic NSCLC (mNSCLC) participants from the Biomarkers of Response to Immunochemotherapy oliGometastatic Hypofractioned radioTherapy study (BRIGHT, NCT04151940). This dual-cohort approach was designed to evaluate the informatics methods generalizability across different treatment response patterns. All participants provided written informed consent prior to enrollment in their respective trial. In both cohorts, all voxel-level analyses were restricted to the primary lung tumor.

#### A.1 FLARE-RT

The FLARE-RT trial received institutional review board approval and was conducted in accordance with our institution’s ethical standards [20]. Between 2016 and 2020, 49 patients with laNSCLC were enrolled and underwent FDG PET/CT imaging and peripheral blood draws prior to treatment initiation and during mid-treatment (week 3) of chemoradiotherapy. Figure 1 (left) shows an example patient with locally advanced disease without distant metastases. For this study, we included the first 25 consecutive patients who had complete mid-treatment PET imaging data.

**Fig. 1.**
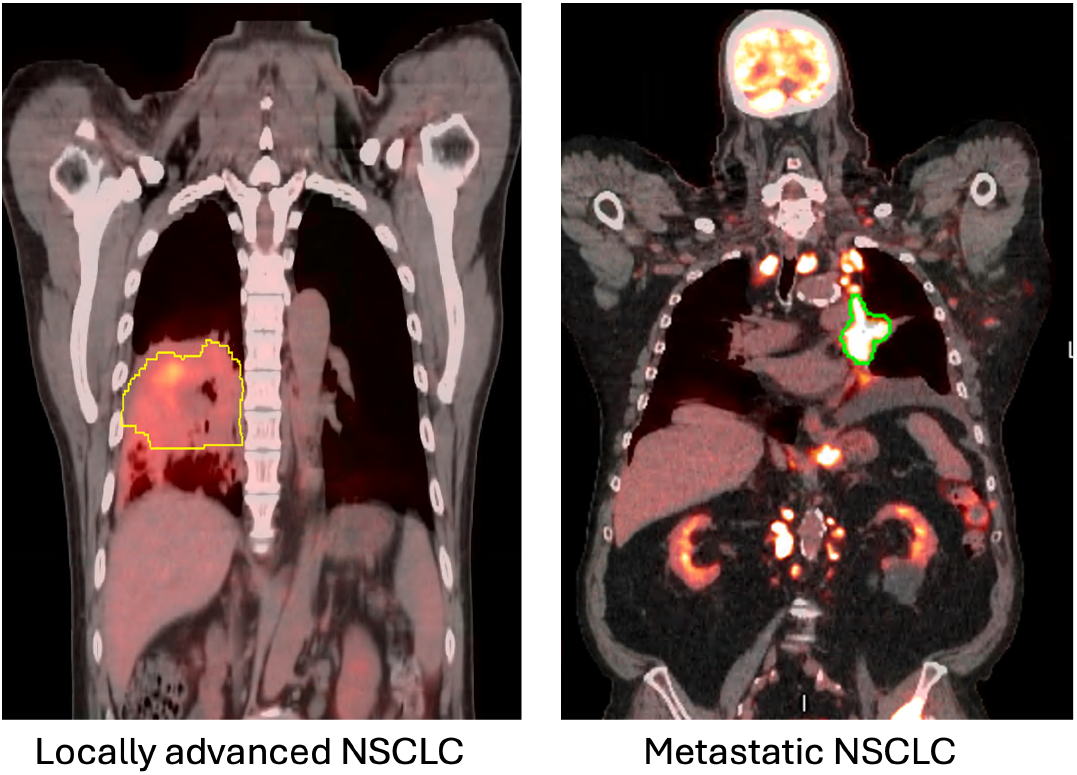
FDG PET/CT of a patient with locally advanced NSCLC (left), showing a bulky primary tumor with high FDG uptake, and a patient with metastatic NSCLC (right).

#### A.2 BRIGHT

The BRIGHT observational study was conducted as part of a prospective clinical trial and received approval from the disease group review, scientific committee, and institutional review board [21], [22]. From 2019 to 2024, 39 patients with mNSCLC (without targetable driver mutations) were enrolled and received first-line carboplatin-pemetrexed-pembrolizumab; a subset additionally received local radiotherapy for oligoprogressive disease. Figure 1 (right) shows an example patient with metastatic disease. Synchronous FDG PET/CT imaging and peripheral blood draws were performed at baseline and after one cycle of combined chemotherapy and immune checkpoint inhibitor (chemoICI) therapy at week 3. The first 19 patients with complete longitudinal imaging data at both time points were included in this study.

### B Data Pre-Processing

#### B.1 FLARE-RT

To preserve the native spatial distribution of FDG PET uptake and minimize uncertainties introduced by image deformation algorithms, alignment in FLARE-RT was restricted to a localized region, and localized rigid registration was applied to the primary tumor. This localized rigid approach ensured preservation of the original voxel geometry within the tumor without introducing deformation-related distortions. Primary tumors were semi-automatically defined on PET/CT using a maximum gradient descent algorithm for PET edge detection (MIM Software Inc.).

#### B.2 BRIGHT

In contrast to the localized rigid alignment used in FLARE-RT, FDG PET/CT images from both time points in BRIGHT were deformably co-registered and resampled onto a common voxel grid for each patient. This deformable registration enabled spatial alignment across multiple regions of interest, including the primary tumor, lungs, liver, and other anatomical structures. Primary tumors were semiautomatically defined using the same gradient-based segmentation tool described above.

### C Proposed Framework

Our proposed methodology is a two-stage framework: In the first stage (1) we utilized our previously developed method called “Voxel Forecast” [13] with re-optimized variogram modeling for spatial correlation in this study and in the second stage (2) we employed our proposed *VarCP* framework to transform point predictions into distribution-free and mathematically guaranteed prediction intervals. The details of these two stages are given below.

#### C.1 Voxel Forecast Framework

We model the midtreatment voxel-level response as a linear mapping between a multiscale predictor space *X* and an observation space *Y* ∈ ℝ^*N*^. The voxel-level response to chemoradiation (FLARERT) and chemoICI (BRIGHT) was defined as the ratio of the FDG PET SUV measured at week 3 divided by the SUV measured at baseline (SUVratio = SUVweek3 / SUVbase-line). Let 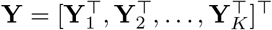 denote the concatenated response vector across *K* independent patients, where each patient *i* contributes *N*_*i*_ voxel-level observations. For patient *i* ∈ 1, …, *K*, the response follows:

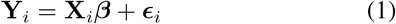

Where 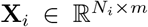 is the multiscale design matrix for patient *i* ∈ {1, …, *K*}, ***β*** ∈ ℝ^*m×*1^ is the vector of regression coefficients (shared across all patients), 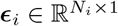 represents spatially-correlated random errors, and 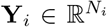 represents voxel-level treatment response (SUV ratio) across *N*_*i*_ voxels indexed by *j*. The design matrix **X**_*i*_ integrates predictors at two hierarchical levels:

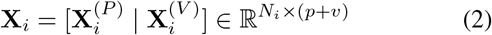

Where 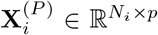 contains patient-level features (e.g., baseline lesion SUV mean; see Table I), constant within patient *i*. Each row of 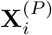 replicates the same *p*-dimensional patientspecific vector across all *N*_*i*_ voxels. 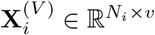 represents voxel-level features (e.g., baseline voxel SUV; see Table I), varying across voxels within patient *i* and *m* = *p* + *v* is the total number of predictors. Correspondingly, the coefficient vector is partitioned as:

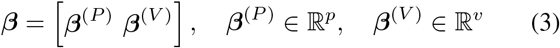

**TABLE I.** Selected Model Features and Their Inclusion in FLARE-RT AND BRIGHT Cohorts.

| Feature | Level | FLARE-RT | BRIGHT |
| --- | --- | --- | --- |
| Intercept | Patient | ✓ | ✓ |
| Stage_iiib (binary) | Patient | ✓ | × |
| log(Lesion SUV mean pre) | Patient | ✓ | ✓ |
| log(Voxel SUV pre) | Voxel | ✓ | ✓ |
| Mid-treatment dose | Voxel | ✓ | × |
| Distance to lesion centroid | Voxel | ✓ | × |

The residual vector 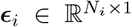 characterizes the unexplained deviations from the deterministic trend. The residual vector ***ϵ***_*i*_ is zero-mean with structured spatial covariance:

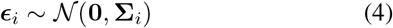

where the patient-specific covariance operator 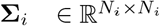 spatial dependency structure of metabolic response. For voxel indices *j, k* ∈ *{*1, …, *N*_*i*_}, the covariance is parameterized as [23]:

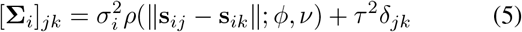

The spatial dependence structure of *ρ*(·) is characterized through the empirical semivariogram, which quantifies how the variance of voxel-level residuals grows with inter-voxel distance [24]:

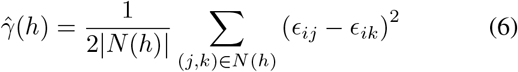

where *N* (*h*) = {(*j, k*)} : ∥**s**_*ij*_ − **s**_*ik*_∥ ≈ *h* is the set of voxel pairs separated by lag distance *h*. Figure 2 illustrates representative empirical semivariograms.

**Fig. 2.**
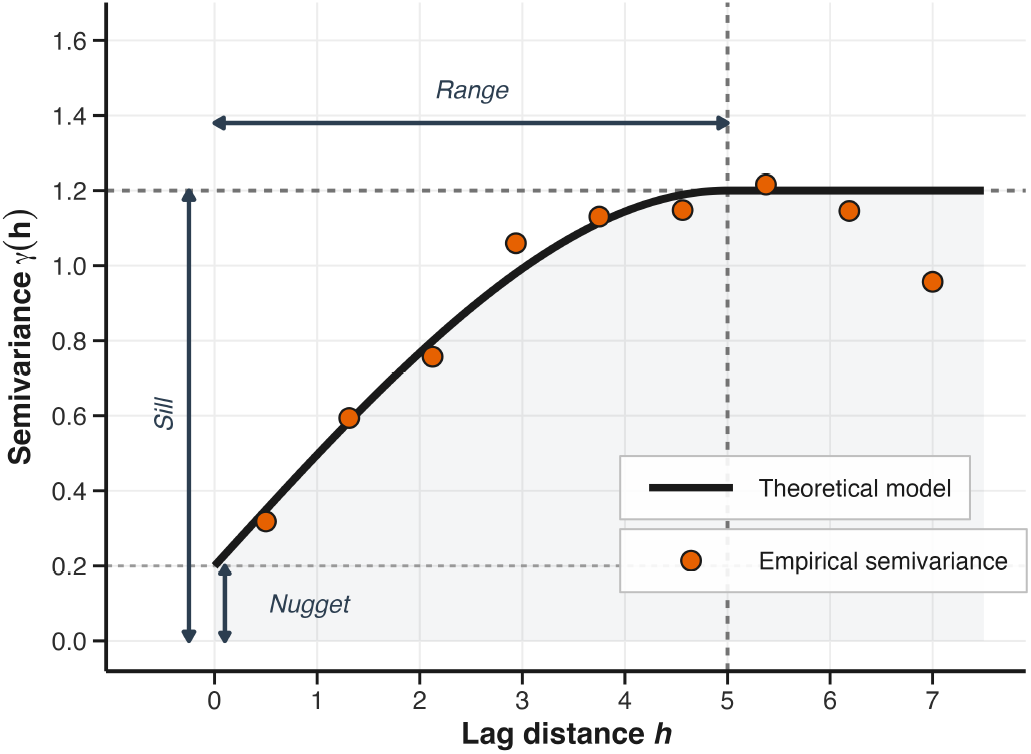
A variogram schema, where orange data points represent the empirical semivariogram. The black curve represents the fitted variogram.

In our previous work [13], *ρ* was the Matérn correlation function. In this work, we evaluated multiple families (see Section C.2 for details) to empirically identify the most robust variogram for voxel-level response prediction based on sensitivity analysis. 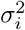 denotes the partial sill (spatial variance component), 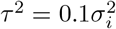 represents the nugget effect accounting for micro-scale metabolic fluctuations, and **s**_*ij*_ ∈ ℝ^3^ are the Cartesian coordinates of voxel *j*. The correlation function *ρ*(·; *ϕ, v*) is parameterized by the effective range *ϕ* and smoothness parameter *v*, while δ_*jk*_ denotes the Kronecker delta.

To stabilize variogram estimation across patients with heterogeneous tumor volumes, we adopt a two-stage procedure. In the first stage, all variogram parameters (sill 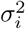, range *ϕ*_*i*_, and smoothness *v*_*i*_) are estimated independently for each patient from the empirical semivariogram. Population-level range and smoothness parameters are then obtained by averaging the patient-specific estimates (geometric mean for range, arithmetic mean for smoothness). In the second stage, patient-specific sills are re-estimated while holding the population-averaged range and smoothness fixed. This yields patient-specific covariance matrices **Σ**_*i*_ defined in equation (4) that share a common correlation decay structure but retain individual variance scaling.

The partitioned structure of ***β*** defined in equation (3) naturally suggests distinct estimation strategies for each component. In standard ordinary least squares (OLS), the estimator minimizes the unweighted residual sum of squares, treating all observations as equally precise and uncorrelated. Generalized least squares (GLS) extends this by incorporating a known covariance structure **Σ**_*i*_, weighting observations inversely by their covariance to yield efficient estimates under spatial correlation.

For patient-level coefficients ***β***^(*P*)^, every voxel within a patient carries the same feature values, so the spatial covariance structure provides no additional information for distinguishing among patients. Consequently, OLS is sufficient and statistically efficient. For voxel-level coefficients ***β***^(*V*)^, however, features vary across spatially correlated voxels, and ignoring this correlation would inflate standard errors and reduce efficiency.

This motivates a hybrid estimation strategy [13] that applies OLS to ***β***^(*P*)^ and GLS to ***β***^(*V*)^, maximizing efficiency at both scales simultaneously. We implement this via block coordinate descent, alternating between two minimization phases until convergence:

##### Phase 1: Update Voxel Coefficients (GLS phase; hold *β*^(*P*)^ fixed)

To account for spatial correlation among voxels within each patient, we update the voxel-level coefficients ***β***^(*V*)^ using GLS. In this step, the patient-level coefficients ***β***^(*P*)^ are held fixed at their current values. We minimize a spatially weighted residual sum of squares, where the weight matrix 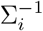 reflects the inverse covariance of voxel errors in patient *i*:

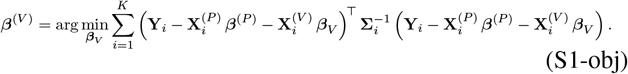

This yields the following closed-form GLS solution:

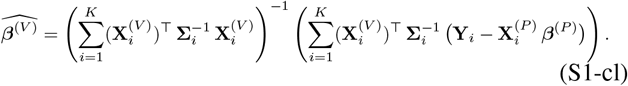

##### Phase 2: Update Patient Coefficients (OLS phase; hold *β*^(*V*)^ fixed)

Next, we update the patient-level coefficients ***β***^(*P*)^, now treating the voxel-level predictions as fixed offsets. Because patient-level features are constant across all voxels within a patient, we use OLS, and minimize the unweighted residual sum of squares:

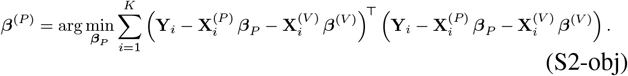

This yields the following OLS estimator:

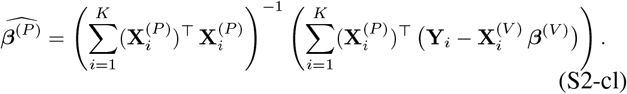

This step treats the voxel-level effects as known offsets and estimates the patient-level trends using standard OLS, which is optimal for patient-invariant features.

##### Convergence criterion

The alternating updates are repeated until ∥***β***^(*t*+1)^ − ***β***^(*t*)^∥_2_ *< ϵ*, where *ϵ* = 10^−4^. Coefficients estimates were summarized using a leave-one-patient-out jackknife to compute standard errors, 95% confidence intervals, and two-sided tests against zero.

While the patient-level update utilizes standard OLS, the voxel-level update (Eq. S1-obj) strictly requires the spatial covariance matrix **Σ**_*i*_ to account for the non-independence of errors within a tumor. Ignoring this correlation would treat spatially clustered residuals as independent data points, leading to invalid inference. To construct **Σ**_*i*_, we model the spatial dependence structure using a variogram, which defines the covariance between any two voxels as a function of their physical distance. Empirically selecting the best-fitting structure is critical for rigorous parameter estimation; therefore, we determined the optimal form via a variogram sensitivity analysis. Parametric variogram models commonly used for spatial correlation include Matérn, Stable, Exponential, Gaussian, and Spherical families, each parameterized by range (*ϕ*) and shape (*v* or *α*) parameters that govern spatial correlation decay.

The features used for the prediction of the voxel-level SUV at mid treatment for both cohorts are shown in Table I

#### C.2 Variogram Model Selection via Sensitivity Analysis

Choosing the right variogram model directly affects the prediction accuracy, as it determines the spatial weighting structure in the generalized least squares estimation [25]. To identify the optimal configuration for voxel-level response prediction, we employed a two-stage variogram model selection strategy as described below.

##### Stage 1: Preliminary Variogram Characterization

In our prior work [26], we evaluated five variogram models including Stable, Matérn, Exponential, Gaussian, and Spherical to model the early response (as ratio) across two independent NSCLC cohorts as mentioned above (FLARE-RT: 25 locally advanced patients; BRIGHT: 19 metastatic patients). Model comparison was based on weighted least squares RMSE between theoretical and empirical semivariograms.

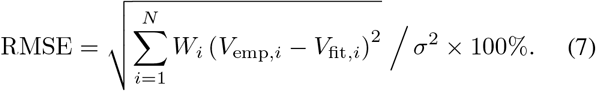

The following prediction-focused sensitivity analysis was conducted using the best performing variogram models out of five (Stable and Matérn).

##### Stage 2: Prediction-Focused Sensitivity Analysis

We conducted a comprehensive sensitivity analysis using leave-one-out cross-validation to test whether superior variogram fit quality translates to improved prediction performance. The following combinations were evaluated:

- Correlation families: Stable, Matérn
- Cutoff percentages: 33%, 50%, 67%, 99% of the maximum pairwise inter-voxel distance, defining the upper lag distance threshold beyond which voxel pairs are excluded from variogram fitting (controlling spatial range estimation)
- Weighting schemes: Weighted vs. unweighted least squares (weighted by bin pair counts during variogram fitting), yielding 16 candidate configurations (2 × 4 × 2) per cohort.

Model performance was compared using leave-one-out cross-validation (LOOCV) metrics, including Mean Absolute Error (MAE), Root Mean Squared Error (RMSE), and their normalized counterparts (MAE ratio and RMSE ratio, divided by the mean observed response), calculated across all patients. Statistical significance of pairwise performance differences was assessed through paired *t*-tests and the non-parametric Wilcoxon signed-rank test with Holm correction for multiple comparisons.

To account for the nested structure of voxels within patients and formally evaluate the independent contribution of each configuration factor while controlling for patient-specific heterogeneity, we utilized a linear mixed-effects model (LMM) defined as:

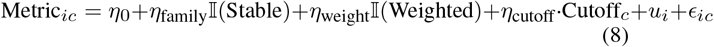

where Metric_*ic*_ is the performance metric (e.g., MAE or RMSE) for patient *i* under configuration *c, η* represents the fixed-effect coefficients for each factor, 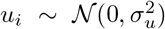 is a patient-level random intercept, and 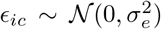 is the residual error. Configuration effects were tested via likelihood ratio tests.

The selected optimal configuration was subsequently used for all downstream prediction tasks, including conformal prediction interval construction.

### D Conformal Prediction Framework

#### D.1 Distribution-Free Prediction Intervals in Log-Space

Conformal prediction (CP) provides distribution-free valid prediction intervals without parametric assumptions on the data-generating process [15]. For a new observation (*X*_new_, *Y*_new_), CP constructs an interval *C*_*α*_(*X*_new_) satisfying the marginal coverage guarantee [15]:

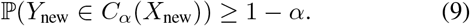

for any miscoverage level *α* ∈ (0, 1). This guarantee relies solely on exchangeability: where a sequence

(*Z*_1_, …, *Z*_*n*_, *Z*_new_) is exchangeable if its joint distribution is invariant under permutations, where *Z*_*i*_ = (*X*_*i*_, *Y*_*i*_). Exchangeability is weaker than the i.i.d. assumption and holds in finite samples, making CP particularly suitable for clinical applications with limited patient cohorts.

The CP framework operates by defining a nonconformity score *R*_*i*_ that quantifies how unusual an observation is relative to the fitted model. For regression, a common choice is the absolute residual, 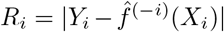, where 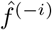 denotes a model trained on all data *except* observation *i*. The prediction interval is then constructed using the empirical (1 − *α*)-quantile of these calibration scores:

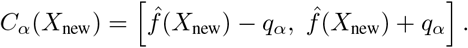

Using 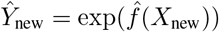, this log-space interval transforms to

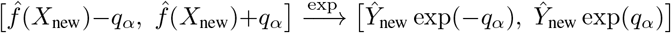

Here *q*_*α*_ = Quantile_1*−α*_({*R*_1_, …, *R*_*n*_}) denotes the smallest *q* such that 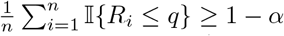.

All models predict in log-space: 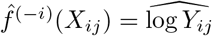, with back-transformation 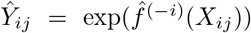. Residuals are computed as:

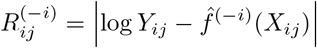

Working in log-space ensures that prediction intervals are multiplicatively symmetric in linear SUV space, respecting the ratio-scale nature of uptake measurements.

#### D.2 Hierarchical Exchangeability

In our voxel-level prediction setting, observations are hierarchically structured: each patient *I* ∈ {1, …, *K*} contains *N*_*i*_ spatially correlated voxels, denoted (*X*_*ij*_, *Y*_*ij*_) for *j* ∈ {1, …, *N*_*i*_}. This hierarchical structure poses a fundamental challenge to the exchangeability assumption required for conformal validity. Naively pooling all voxels as independent observations would violate exchangeability, since Cov(*Y*_*ij*_, *Y*_*ik*_ | *X*_*i*_) = [**Σ**_*i*_]_*jk*_ ≠ 0 for *j* ≠ *k*.

We assume that patients are i.i.d. (or, more generally, exchangeable) draws from the underlying population, while voxels within a given patient may be arbitrarily dependent. We restore validity by treating each patient as an exchangeable block 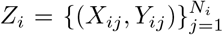, exploiting the fact that patients are independent samples from the population. Under patient-level exchangeability, the marginal voxel distribution is:

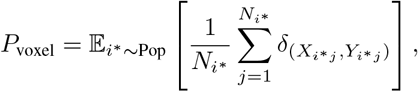

where δ_(*x,y*)_ is the Dirac measure at (*x, y*). Because *P*_voxel_ is invariant to permutations of the patient blocks, pooling voxel-wise scores across all patients preserves the block-wise exchangeability needed for conformal validity.

The pooled-voxel coverage guarantee we establish in Theorem 1 below corresponds to the probability that a voxel drawn uniformly at random from the entire cohort, i.e., uniformly over patients and voxels within each patient, is covered by the prediction interval.

#### D.3 Nested Leave-One-Out Calibration

To construct valid intervals while respecting the hierarchical structure, we employ a nested leave-one-out (LOO) procedure. For each patient *i* ∈ {1, …, *K*}:

- Train a model 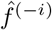 on all patients except patient *i*.
- Compute voxel-level nonconformity scores within patient *i*:

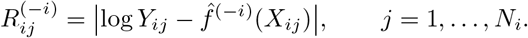
- Form the patient-specific score block

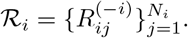

The pooled calibration set is then the union of these patient blocks:

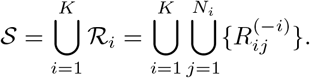

Under patient-level exchangeability, the score blocks

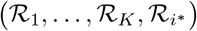

are jointly exchangeable as blocks, where 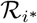 denotes the (unobserved) score block that would be obtained for a new test patient using the corresponding leave-one-out model 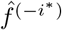. This block-exchangeability formalizes that calibration scores for any test patient *i*^*\**^ are drawn from the same joint distribution as the observed blocks and are always computed by models that never trained on patient *i*^*\**^. Consequently, standard conformal prediction arguments for grouped (block-structured) data apply directly when we pool scores across patients using *S*.

#### D.4 Standard Conformal Prediction (StdCP)

The baseline method is standard (homoscedastic) conformal prediction, which uses a single global residual quantile. Given the pooled calibration set *S*, the (1 − *α*)-quantile

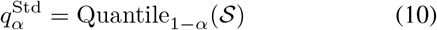

defines the half-width of the log-space prediction interval.

For a test patient *i*^*\**^, we train 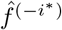 on all remaining patients. For each voxel *j* *

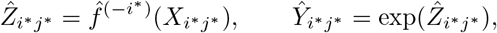

and the standard conformal interval in SUV space is

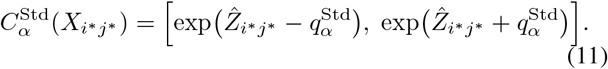

This interval has constant multiplicative width across voxels, determined entirely by the global distribution of calibration residuals.

#### D.5 Residual-Variance Conformal Prediction (VarCP)

Standard CP treats all voxels as having the same residual scale, which can be suboptimal when prediction difficulty varies substantially across the tumor volume. To allow voxel-level intervals to adapt to predicted local difficulty, we introduce a residual-variance conformal prediction (VarCP) method that normalizes residuals by a learned heteroscedastic scale model.

Using the absolute log-residuals 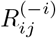 defined in the nested LOO procedure, we train a regression model *g* (Random Forest with 200 trees and maximum depth of 6 to regularize the variance estimate) from the same multiscale features used in the Voxel Forecast to predict their magnitude,

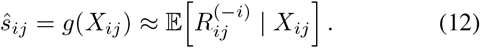

To stabilize the scale and avoid overly aggressive shrinkage in regions where *ŝ*_*ij*_ is small, we add a constant *c* equal to the median predicted scale over all calibration voxels,

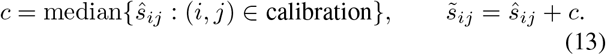

The normalized nonconformity score for voxel (*i, j*) is then

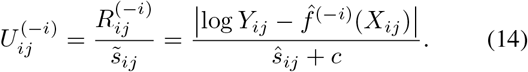

Pooling these scores across all patients yields

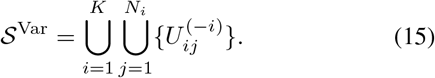

The (1 − *α*)-quantile of the normalized scores,

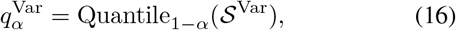

defines the global calibration constant for VarCP.

For a new voxel (*i*^*\**^, *j*^*\**^), we compute the out-of-sample prediction and scale

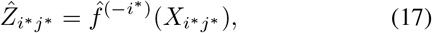

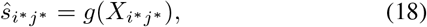

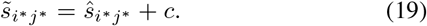

The VarCP log-space half-width is

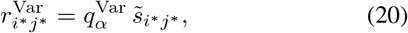

and the corresponding SUV-space interval is

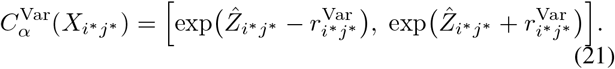

Compared to Standard CP, VarCP allows intervals to be shorter in regions where the residual-variance model predicts low difficulty and wider where it predicts high difficulty, while using a single global quantile on the normalized scores.

#### D.6 Coverage Guarantees

Both Standard CP and VarCP fit within the same conformal framework; they differ only in the choice of nonconformity score (unnormalized vs. normalized). Under patient-level exchangeability, we obtain the following marginal voxel coverage guarantee.

##### Theorem 1

(Marginal Voxel Coverage for StdCP and VarCP). *Under patient-level exchangeability and the nested leave-one-out construction described above, both the Standard CP interval* 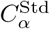 *in* (11) *and the VarCP interval* 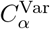 *in* (21)

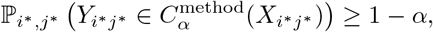

*where* method ∈ {Std, Var} *and* ℙ_*i*, j\**_ *denotes the probability when* (*i*^*\**^, *j*^*\**^) *is drawn by first sampling a patient i*^*\**^ *from the population and then sampling a voxel j*^*\**^ *uniformly from* 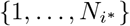.

*Proof*

This result follows directly from standard conformal prediction guarantee under block-exchangeability [27] applied to our nested leave-one-out construction. For each method, the scores 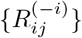 (Standard CP) or 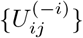 (VarCP) are computed in a leave-one-patient-out fashion and pooled across patients. Under patient-level exchangeability, the corresponding patient score blocks are jointly exchangeable, so the rank of any test score among all calibration scores and the test score is uniform. Since 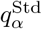 and 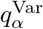 are chosen so that at least a (1 − *α*) fraction of calibration scores are at or below the threshold, the usual conformal argument [15], [27] implies ℙ (test score 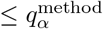) ≥ 1 − *α*, which is equivalent to the stated coverage property in log-space and hence in SUV space.

An immediate corollary is a guarantee on the expected fraction of covered voxels within a randomly selected test patient.

##### Corollary 1

(Expected Voxel-Wise Coverage per Patient). *Under patient-level exchangeability, the expected fraction of covered voxels within a randomly selected test patient satisfies:*

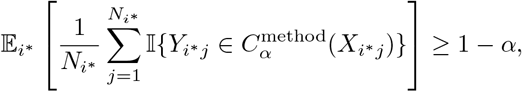

*for both Standard CP and VarCP*.

*Proof:* Follows from Theorem 1 by the law of iterated expectations [28], [29].

[Scope of the formal guarantee] Theorem 1 and Corollary 1 establish marginal coverage guarantees at the *voxel level* only. To evaluate clinical utility at a coarser scale, we additionally assess lesion-level coverage, in which each patient’s primary tumor is treated as a single lesion: the lesion-level prediction interval is derived by taking the median of the voxel-level lower and upper bounds, and coverage is defined as whether the true lesion-mean SUV ratio falls within this aggregated interval. This lesion-level summary does not inherit a formal distribution-free guarantee and is therefore reported as an empirical observation rather than a theoretically guaranteed coverage property.

#### D.7 Overall Algorithm

The full analysis pipeline combines the Voxel Forecast with both standard CP (StdCP) and residual-variance CP (VarCP). Stage A fits the Voxel Forecast model using a Stable variogram and hybrid GLS/OLS estimation. Stage B runs a nested leave-one-patient-out loop to compute out-of-sample voxel-level residuals, fits a Random Forest residual-variance model *σ*(*x*) on these residuals, and computes the empirical quantiles for StdCP and VarCP. Stage C applies these quantiles to construct voxel-level prediction intervals for held-out test patients under both methods and aggregates voxel-level metrics to the lesion and patient levels.

#### D.8 Experimental setup

All analyses were conducted independently on each cohort using nested leave-one-out cross-validation. The experimental pipeline proceeded in three stages. First, a variogram sensitivity analysis was performed to identify the optimal spatial correlation structure for the Voxel-Forecast model. Second, the selected configuration was used to evaluate multi-scale prediction accuracy by comparing a patient-level-only model against the full patient + voxel-level model. Third, the residuals from the multi-scale model were used to construct and evaluate both StdCP and VarCP intervals across three miscoverage levels (*α* = 0.10, 0.15, 0.20, corresponding to nominal 90%, 85%, and 80% two-sided prediction intervals), assessing coverage validity, and interval width. These levels were selected to balance clinical interpretability with quantile stability, as tighter levels such as *α* = 0.05 would rely on extreme quantile estimates with high variance.

All computational analyses, including the estimation of large spatial covariance matrices via Cholesky decomposition [30] during generalized least squares optimization, were implemented in Python. The nested leave-one-patient-out conformal calibration procedure, which requires training *O*(*K*^2^) models for *K* patients, was parallelized across multiple CPU cores to accelerate execution. All experiments were conducted on an Apple MacBook Pro (M3 Max chip, 64 GB RAM) without GPU acceleration, demonstrating the computational feasibility of the framework on standard high-performance consumer hardware.

## III Results

### A Patient and Tumor Characteristics

Table II summarizes descriptive cohort characteristics of patient and tumor (not model inputs; see Table I for features used in the predictive model) for both FLARE-RT and BRIGHT cohorts. Primary tumor voxels were *n*_vox_ = 11,100 for FLARE-RT and *n*_vox_ = 26,980 for BRIGHT. BRIGHT primary tumors were substantially larger (metabolic tumor volume [MTV]: 101 vs. 27 cm^3^) and more metabolically heterogeneous (coefficient of variation [CV]: 0.50 vs. 0.32) than FLARE-RT tumors. The response ratio (mid-treatment lesion-mean SUV / baseline lesion-mean SUV) was lower in FLARE-RT (0.62 vs. 0.78), reflecting deeper and more consistent metabolic suppression, in line with the pronounced dose-response relationship in radiotherapy.

**TABLE II.** Patient and Tumor Characteristics.

| Characteristic | FLARE-RT<br>(*n* = 25) | BRIGHT<br>(*n* = 19) |
| --- | --- | --- |
| Age (years) | 62 (50–75) | 65 (41–84) |
| **Sex** |  |  |
| Female | 15 (60%) | 12 (63%) |
| Male | 10 (40%) | 7 (37%) |
| **Smoking History** |  |  |
| Never | 5 (20%) | 5 (26%) |
| Former | 18 (72%) | 8 (42%) |
| Current | 2 (8%) | 6 (32%) |
| **Primary Tumor Imaging** |  |  |
| MTV (cm<sup>3</sup>) | 27 (2–396) | 101 (7–609) |
| Baseline SUV<sub>mean</sub> | 6 (2–16) | 4 (2–8) |
| Baseline SUV<sub>max</sub> | 11 (3–31) | 12 (4–26) |
| CV (heterogeneity) | 0.32 (0.18–0.48) | 0.50 (0.29–0.90) |
| Lesion size (voxels) | 407 (146–796) | 1032 (348–3817) |
| **Treatment Response** |  |  |
| Response ratio (mid/pre) | 0.62 (0.27–1.30) | 0.78 (0.37–1.40) |
| Ratio < 1.0 | 23 (92%) | 15 (79%) |
| Ratio ≥ 1.0 | 2 (8%) | 4 (21%) |
| **Voxels Analyzed** | 11,100 | 26,980 |

Values are median (min–max) for continuous variables and *n* (%) for categorical variables. MTV = metabolic tumor volume; CV = coefficient of variation (SD/mean) of baseline voxel SUV, a marker of intra-tumoral heterogeneity. Response defined as mid-treatment lesion-mean SUV/baseline lesion-mean SUV < 1.0.

#### Algorithm 1

Voxel Forecast + StdCP/VarCP

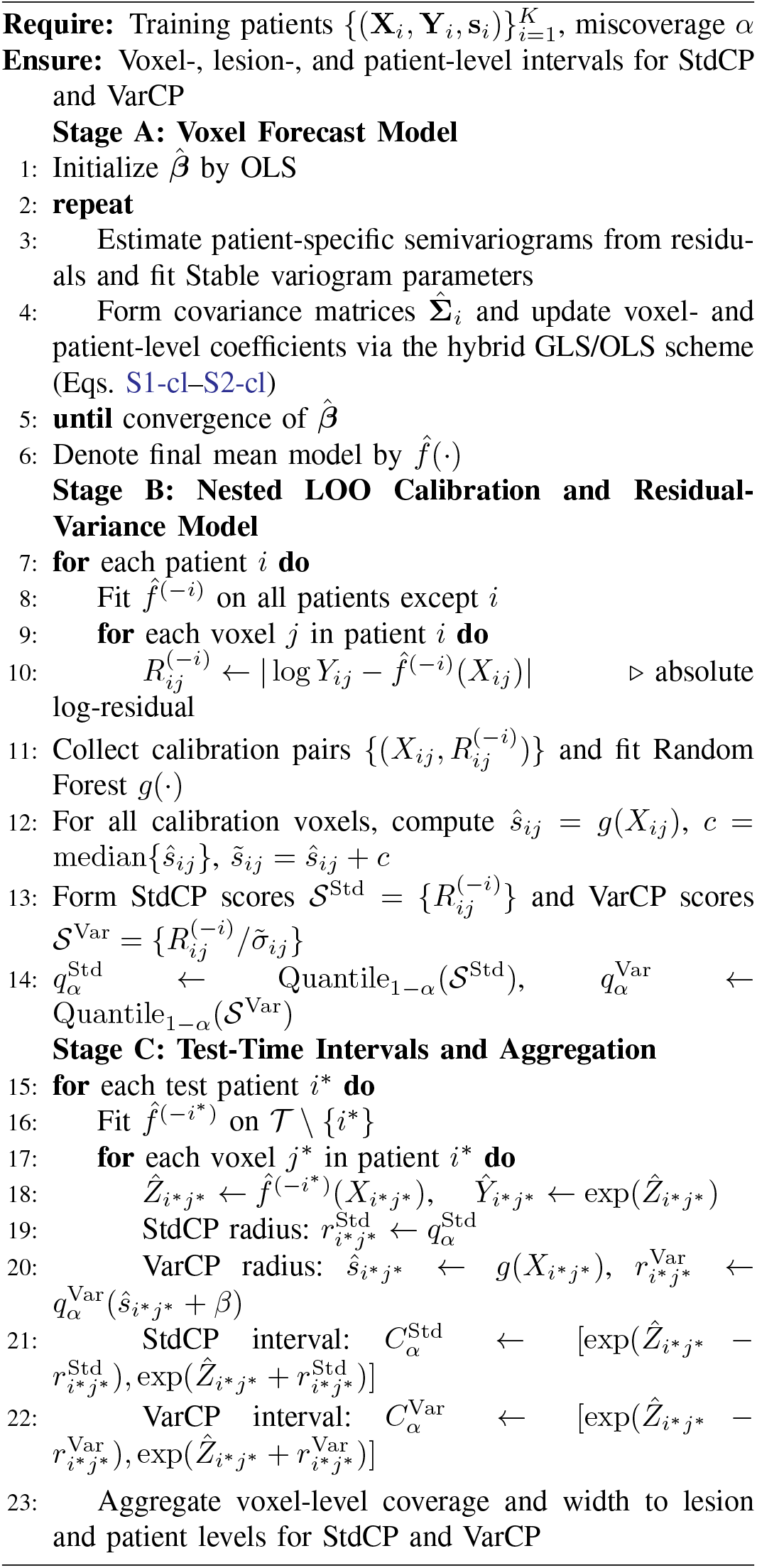

### Variogram selection and optimization

#### B.1 Variogram selection

Supplementary Figure 1 shows the boxplots of the RMSE for different variogram models across FLARE-RT and BRIGHT. The Stable model achieved the lowest root mean squared error (RMSE) on average (mean: 5.2-5.5%), followed by the Matérn model (mean: 5.8-7.4%), both of which performed better than most other models. In contrast, the Exponential model had the highest RMSE (mean: 9.3-15.6%).

#### B.2 Variogram optimization

Supplementary Table 1 summarizes the sensitivity analysis across 16 configurations. The *Stable* variogram family yielded significantly lower prediction errors than *Matérn* across all metrics in both cohorts (BRIGHT: *β* = −0.019, 95% CI [−0.021, −0.018], *p <* 0.001; FLARE-RT: *β* = −0.006, 95% CI [−0.007, −0.005], *p <* 0.001 for RMSE). The weighting scheme had no significant effect in BRIGHT (*p >* 0.90), while unweighted predictions were significantly better in FLARE-RT (*β* = 0.004, *p <* 0.001 for RMSE). The cutoff percentage had a negligible effect on performance (*β* ≈ − 3 × 10^−5^ per percentage point). These findings were corroborated by Wilcoxon signed-rank tests with Holm correction. Based on these results, the Stable family with 99% cutoff and an unweighted scheme was adopted as the final configuration

### C Multi-Scale Features Improve Prediction

Incorporating voxel-level spatial features alongside patient-level features (Table I) significantly reduced the relative prediction error in both cohorts (FLARE-RT: RMSE ratio = 0.22 vs. 0.34, *p <* 0.05; BRIGHT: 0.44 vs. 0.64, *p <* 0.05; Table III). This demonstrates that multi-scale feature integration captures complementary information critical for voxel-level response modelling, providing a robust foundation for subsequent uncertainty quantification. This difference in features (Table I) and treatment regime, combined with the higher intra-tumoral heterogeneity of BRIGHT primary tumors (CV = 0.50 vs. 0.32), accounts for the larger residual error in BRIGHT (RMSE ratio 0.44 vs. 0.22), and motivates the adaptive uncertainty quantification described below. Predictors that remained statistically significant (*p <* 0.05) across models and cohorts were baseline lesion SUV_mean_ (Model: Patient-level) and baseline voxel SUV (Model:Patient+ voxel-level), as summarized in Supplementary Table 3.

**TABLE III.** Impact of Voxel-Level Features on Prediction Accuracy (Relative RMSE)

| Dataset | Modeling Approach | Relative RMSE | Improvement |
| --- | --- | --- | --- |
| FLARE-RT | Patient-level | 0.34 | — |
| FLARE-RT | Patient + voxel-level | **0.22** | ↓35% |
| BRIGHT | Patient-level | 0.64 | — |
| BRIGHT | Patient + voxel-level | **0.44** | ↓31% |

### D Conformal Prediction

#### D.1 Voxel-level coverage

At the voxel level, both StdCP and VarCP achieved coverage close to the nominal (1 − *α*) targets in FLARE-RT and BRIGHT (Table IV). In FLARE-RT, VarCP yielded consistently narrower intervals than StdCP at all three miscoverage levels while maintaining nearly identical coverage (differences ≤ 0.003). The mean voxel-level width was reduced by 7.0%–17.9% across all *α*, with all reductions reaching statistical significance on a per-patient basis (paired Wilcoxon signed-rank test). In BRIGHT, VarCP again produced numerically narrower intervals than StdCP (3.8–8.1% mean width reduction across *α*), with essentially unchanged or slightly higher coverage, although the paired width reduction reached conventional significance only at *α* = 0.10 (*p <* 0.05) and trended toward significance at *α* = 0.15 (*p* ≈ 0.06), consistent with the smaller cohort size and greater inter-patient heterogeneity.

**TABLE IV.** Voxel-level coverage and interval width for Standard CP (StdCP) AND RESIDUAL-VARIANCE CP (VarCP).

| Cohort | $\alpha$ | Coverage<br>Mean ± SEM | | Width<br>Mean ± SD | | Rel. gain (%) | $p_{\text{width}}$ |
| --- | --- | --- | --- | --- | --- | --- | --- |
|  |  | StdCP | VarCP | StdCP | VarCP |  |  |
| FLARE-RT | 0.10 | 0.917 ± 0.030 | 0.914 ± 0.017 | 5.42 ± 1.60 | 4.45 ± 2.09 | 17.9 | $p < 0.001$ |
| | 0.15 | 0.872 ± 0.038 | 0.869 ± 0.024 | 4.53 ± 1.34 | 3.98 ± 1.85 | 12.1 | $p < 0.01$ |
| | 0.20 | 0.826 ± 0.043 | 0.823 ± 0.030 | 3.91 ± 1.16 | 3.64 ± 1.67 | 7.0 | $p < 0.05$ |
| BRIGHT | 0.10 | 0.901 ± 0.021 | 0.903 ± 0.013 | 4.42 ± 0.83 | 4.06 ± 1.07 | 8.1 | $p < 0.05$ |
| | 0.15 | 0.843 ± 0.028 | 0.845 ± 0.017 | 3.72 ± 0.70 | 3.47 ± 0.90 | 6.7 | $p = 0.06$ |
| | 0.20 | 0.787 ± 0.035 | 0.791 ± 0.021 | 3.21 ± 0.60 | 3.08 ± 0.79 | 3.8 | n.s. |

Coverage values are mean ± SEM across patients. Width values are mean ± SD across patients. Relative gain = (StdCP width - VarCP width)/StdCP width. $p_{\text{width}}$ from paired Wilcoxon signed-rank test on per-patient mean widths (one-sided, StdCP > VarCP).

#### D.2 Lesion-level coverage

In both cohorts, empirical lesion-level coverage was at or above the nominal (1 − *α*) target for both methods (Table V; note that lesion-level coverage carries no formal guarantee and is reported as an empirical finding). VarCP yielded narrower lesion-level intervals than StdCP at all three miscoverage levels, with relative width reductions of 18% (FLARE-RT) and 10% (BRIGHT) at *α* = 0.10. The difference in interval width was statistically significant (paired Wilcoxon signed-rank test, VarCP narrower than StdCP) in FLARE-RT at all *α* (*p* ≤ 0.02) and in BRIGHT at *α* = 0.10 and 0.15 (*p* ≤ 0.02); at *α* = 0.20 in BRIGHT the reduction remained positive but was not significant (*p* = 0.06). These results indicate that the efficiency gains of VarCP at the voxel level carry over to the lesion level when intervals are aggregated via the median, supporting the use of VarCP for both voxel-wise and whole-lesion uncertainty quantification.

**TABLE V.** Lesion-level coverage and interval width for Standard CP (StdCP) and residual-variance CP (VarCP)

| Cohort | $\alpha$ | Coverage (Mean ± SEM) | | Width (Mean ± SD) | | Rel. gain | |
| --- | --- | --- | --- | --- | --- | --- | --- |
| | | StdCP | VarCP | StdCP | VarCP | (%) | $p_{\text{width}}$ |
| FLARE-RT | 0.10 | 1.000 ± 0.000 | 1.000 ± 0.000 | 5.25 ± 1.57 | 4.30 ± 2.02 | 18.1, $p < 0.001$ | |
| | 0.15 | 0.920 ± 0.055 | 1.000 ± 0.000 | 4.39 ± 1.32 | 3.85 ± 1.80 | 12.2, $p < 0.01$ | |
| | 0.20 | 0.920 ± 0.055 | 1.000 ± 0.000 | 3.79 ± 1.14 | 3.52 ± 1.63 | 7.1, $p < 0.05$ | |
| BRIGHT | 0.10 | 1.000 ± 0.000 | 1.000 ± 0.000 | 4.36 ± 0.83 | 3.93 ± 1.09 | 9.8, $p < 0.01$ | |
| | 0.15 | 1.000 ± 0.000 | 1.000 ± 0.000 | 3.68 ± 0.71 | 3.37 ± 0.92 | 8.4, $p < 0.05$ | |
| | 0.20 | 1.000 ± 0.000 | 1.000 ± 0.000 | 3.16 ± 0.60 | 2.98 ± 0.81 | 5.6, $p = 0.06$ | |

Coverage values are mean ± SEM across patients. Width values are mean ± SD across patients. Relative gain = (StdCP width - VarCP width)/StdCP width. $p_{\text{width}}$ from paired Wilcoxon signed-rank test on per-patient mean widths (one-sided, StdCP > VarCP).

#### D.3 Qualitative examples

Figure 3 illustrates four representative patients selected to demonstrate the clinical utility of our multiscale framework across both cohorts at *α* = 0.20. This supports personalized voxel-level dose-adjustment by providing rigorous prediction intervals that capture spatial heterogeneity in treatment response, enabling risk-aware dose-adjustments at both voxel and lesion levels. Voxel-level intervals enable voxel-specific dose-adjustment by identifying regions with high confidence in response prediction (green: within interval) versus regions with greater uncertainty (red: outside interval). Lesion-level intervals (Fig. 3, right)support whole-lesion treatment decisions by aggregating voxel-level information using median-based calibration. The multiscale approach demonstrates synergistic value: for example, the qualitative examples (BRIGHT Patient 7 and FLARE-RT Patient 10) exhibit substantially poorer voxel-level coverage under VarCP, while their lesion-level intervals remain covered. This highlights how voxel-level uncertainty maps provide a critical safety measure by revealing spatial heterogeneity and localized regions of uncertainty that may be masked by lesion-level aggregation. This synergy allows clinicians to leverage lesion-level robustness for treatment planning while using voxel-level granularity as a safety check to identify subregions requiring more conservative interpretation despite favorable lesion-level predictions.

**Fig. 3.**
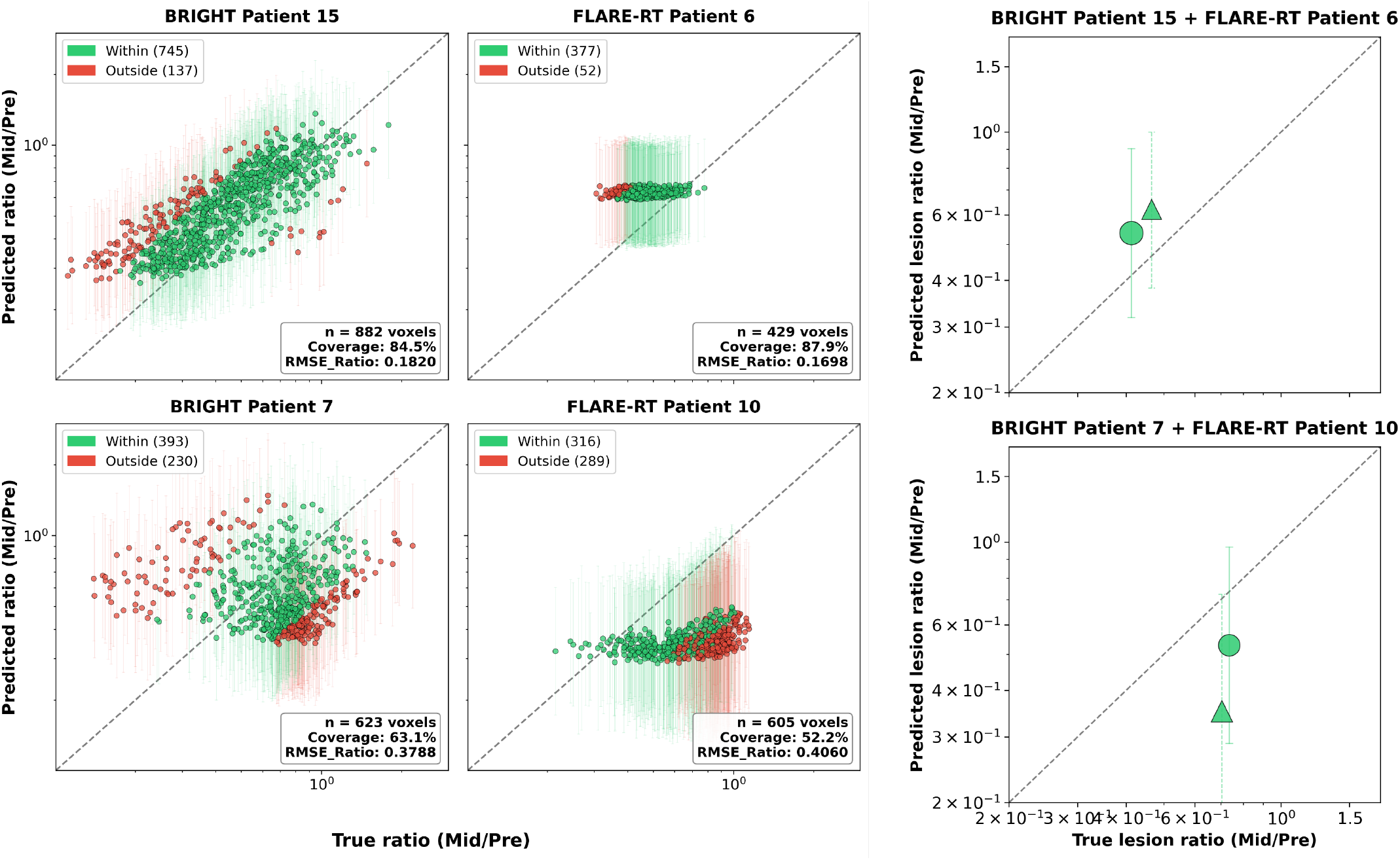
Representative best-(top row) and worst-covered (bottom row) patients from each cohort at ***α* = 0.2**. Voxel-level panels show true vs. predicted mid/pre SUV ratio (green = within interval; red = outside). Lesion-level panel shows predicted lesion-ratio intervals with coverage status(circles: BRIGHT, triangles: FLARE-RT; green: lesion median in interval, red: outside). Annotations include voxel count, voxel coverage, mean interval width ***±*** SD, and RMSE ratio. All plots use logarithmic axes.

In both cohorts, VarCP out-of-interval voxels (red) were not spatially random within the primary tumor MTV. In BRIGHT (*α* = 0.20), uncovered voxels were significantly more peripheral than covered voxels (median centroid distance 2.60 vs. 2.45; Mann–Whitney *p <* 0.001) and had higher baseline uptake (median SUV_pre_ 3.45 vs. 2.98; *p <* 0.001). FLARE-RT showed the same pattern, with uncovered voxels occurring farther from the centroid (2.06 vs. 1.59; *p <* 0.001) and at higher baseline SUV_pre_ (6.92 vs. 5.42; *p <* 0.001). These results support the qualitative examples by showing that VarCP failures concentrate in peripheral, high-uptake regions that are plausibly harder to predict.

VarCP interval width provides a practical reliability signal: intervals are tightest when the predicted residual variance is low and widen automatically when response is harder to forecast. Across both cohorts (*α* = 0.20), mean VarCP width increased with baseline uptake and the learned residual scale *ŝ* (*x*). Specifically, VarCP width was strongly and positively correlated with baseline SUV_pre_ (FLARE-RT: *ρ* = 0.91, *p <* 0.001; BRIGHT: *ρ* = 0.70, *p <* 0.001); note that part of this association is inherent to the multiplicative back-transformation from log-space, whereby higher predicted responses yield wider linear-space intervals even at fixed log-space half-width. VarCP width was additionally correlated with the learned scale *ŝ*(*x*) (FLARE-RT: *ρ* = 0.59, *p <* 0.001; BRIGHT: *ρ* = 0.72, *p <* 0.001), confirming that the scale model captures genuine spatial heterogeneity in prediction difficulty beyond this structural effect. Thus, the model is most reliable (tighter intervals) in lower-uptake tumors and regions with low predicted residual scale, and least reliable (wider intervals) in higher-uptake, higher-uncertainty regions.

## IV Discussion

### A Summary

Building on a systematic variogram sensitivity analysis that identified the Stable model as optimal for voxel-level response prediction, this study introduced VarCP, a residual-variance conformal prediction framework for voxel-level uncertainty quantification. This framework augments standard conformal prediction by learning a residual-variance model to construct heteroscedastic, distribution-free prediction intervals while preserving marginal coverage guarantees. Through a multi-scale evaluation, we showed how these voxel-level prediction intervals aggregate via median-based lesion summaries to yield empirical lesion-level coverage that supports both voxel-level dose-adjustment and whole-tumor treatment decisions. Finally, we validated the approach across two independent prospective NSCLC trial cohorts with distinct disease stages and treatment paradigms, demonstrating its robustness across heterogeneous clinical settings.

### B Variogram

A two-stage analysis, comprising initial fit-quality selection from five candidate families (Stable, Matérn, Exponential, Gaussian, Spherical) followed by prediction-focused sensitivity analysis on the two top-performing families, identified the Stable (powered exponential) model as significantly superior to Matérn (*p <* 0.001) across both cohorts. While the geostatistical literature has traditionally favored the Matérn family for its theoretical flexibility [23], [24], [31], our results demonstrate that the Stable model offers a robust empirical alternative for characterizing heterogeneous tumor response on FDG-PET imaging. These findings indicate that the Stable model’s additional shape parameter provides the flexibility needed to capture the sharp spatial transitions and high-gradient microen-vironmental shifts characteristic of tumor boundaries. This is consistent with prior findings that, when a spatial field is non-differentiable or rapidly varying, the Powered Exponential class can offer a superior empirical fit relative to standard Matérn parameterizations [32]–[34].

### C Voxel Forecast

Our results show that incorporating voxel-level features alongside patient-level features significantly improves prediction in both cohorts (*p <* 0.001), reducing the RMSE ratio by 35 % in FLARE-RT and 31 % in BRIGHT. The three patient-level features (Table I) were retained from our prior work [13], where they were pre-selected as statistically significant predictors of voxel-level response. These findings are consistent with our prior work [13] and align with the broader literature on predictive modeling. For example, *Escobar et al*. [35] demonstrated that voxel-wise supervised radiomic features from PET/CT can identify predictive biological patterns and improve patient stratification in soft-tissue sarcoma, underscoring the value of preserving spatial resolution over conventional ROI-level analyses. Similarly, *Zhang et al*. [36] showed that voxel-level radiomics combined with deep learning provides more detailed and robust tumor-response prediction than region-based methods, with consistent improvements across multiple independent validation cohorts. The performance gap between cohorts (RMSE ratio 0.22 vs. 0.44) likely reflects BRIGHT’s greater intratumoral metabolic heterogeneity (CV 0.50 vs. 0.32), distinct treatment regimens (FLARE-RT: chemoradiation vs. BRIGHT: chemoimmunotherapy), and the smaller BRIGHT cohort (*n* = 19 vs. *n* = 25), which constrains the model’s ability to learn from more limited patient-level variability.

### D Conformal Prediction Results

Across all six evaluation settings (2 cohorts × 3 *α* levels), the empirical coverage of VarCP remained close to the nominal (1 − *α*) targets and closely tracked standard conformal prediction (StdCP), consistent with the finite-sample coverage guarantees of conformal prediction under exchangeability [15], [37]. FLARE-RT yielded narrower SUV intervals than BRIGHT (reflecting its lower prediction residuals), and VarCP consistently produced shorter voxel-level intervals than StdCP in both cohorts (Table IV). This tight coupling between point-prediction accuracy, residual concentration, and interval width is intrinsic to conformal regression [15], [38]: when residuals are more concentrated or better explained by the learned variance model *σ*(*x*), the calibration quantiles of the normalized nonconformity scores decrease, producing shorter yet still approximately valid prediction intervals. VarCP achieved coverage closely aligned with StdCP at all evaluated miscoverage levels in both cohorts, while yielding systematically narrower intervals. In FLARE-RT, VarCP reduced mean voxel-level clinical SUV interval width by approximately 7–18% relative to StdCP, with smaller but consistent gains in BRIGHT, and these reductions were statistically significant at most *α* levels (Table IV). This pattern is consistent with the behavior of distribution-free conformal methods [15], [39], which are designed to guarantee finite-sample coverage under weak assumptions and therefore cannot aggressively shrink intervals without sacrificing this guarantee. Within this constraint, adaptive conformal methods have been shown to deliver modest but meaningful efficiency gains in both regression and classification by leveraging covariate- or feature-dependent variability [40]–[42]. Our results, showing absolute reductions on the order of 0.13–0.97 SUV units (corresponding to roughly 4–18% relative gains) at the voxel-level, align with these prior findings and demonstrate that a residual-variance–based VarCP can provide tighter yet still reliable prediction intervals for FDG-PET treatment response. Importantly, these efficiency gains translated to the lesion scale: VarCP maintained lesion-level coverage at or above target in both cohorts while reducing mean lesion interval width by roughly 7–18% in FLARE-RT and 5–10% in BRIGHT, with significant paired reductions at *α* = 0.10 and 0.15 (Table V).

### E Limitations

Several limitations need to be acknowledged. The proposed method Voxel Forecast and VarCP framework depends on precisely registered FDG PET/CT and accurate metabolic tumor volume delineation as inputs; inter-observer variability in manual contouring or misregistration artifacts would propagate directly into the variogram estimation and spatial covariance matrix, subsequently inflating prediction errors and ultimately widening the resulting conformal prediction intervals. Second, although our analysis leverages tens of thousands of voxels cumulatively, both cohorts are relatively small in terms of distinct patients (FLARE-RT: *n* = 25, BRIGHT: *n* = 19). A fundamental advantage of conformal prediction is that its marginal coverage guarantees remain mathematically valid in finite samples, regardless of cohort size [15]; however, smaller patient pools naturally increase the variance of the empirical quantile estimates. Consequently, larger datasets would likely yield tighter and more stable conformal intervals, and would also enable formal assessment of how coverage varies across patient subgroups (e.g., early versus late responders), which was not feasible with the current sample size. Finally, the current study does not provide an end-to-end software tool for clinician interaction. While validating the Voxel Forecast and core uncertainty quantification framework (VarCP) was the primary objective of this work, the development and evaluation of a clinician-facing tool using human computer interaction (HCI) principles and clinician feedback were not included in the current study.

### F Future work

We plan to validate our proposed Voxel Forecast and VarCP framework on larger datasets as enrollment in the BRIGHT clinical trial is still ongoing. Second, while our current analysis focused exclusively on primary NSCLC tumors, assessing the spatial correlation structures and conformal prediction framework for metastatic lesions remains a critical next step. Third, we aim to investigate the generalizability of our re-optimized Voxel-Forecast and conformal prediction framework by applying it to other solid tumor types where spatially heterogeneous treatment response is clinically actionable. Finally, addressing the lack of an interactive clinical interface in the current study, a crucial direction for clinical translation is the design and prospective evaluation of a multilayered visualization tool that seamlessly integrates voxel-level confidence maps with lesion-level summary data, empowering physicians to interactively balance macroscopic efficacy with microscopic uncertainty during adaptive treatment planning.

### G Conclusion

This study established a unified framework for uncertainty-aware multiscale treatment response prediction in advanced NSCLC. By conducting a systematic sensitivity analysis, we first identified the Stable variogram as the robust and optimal model for spatial correlation, significantly improving voxel-level predictions. We then introduced VarCP, a residual-variance conformal prediction framework that provides rigorous, distribution-free uncertainty quantification by learning a variance model to adapt interval width to local prediction difficulty while preserving marginal coverage guarantees.

Our multiscale evaluation verified that these guaranteed voxel-level intervals are tighter on average than standard conformal prediction, thereby providing greater certainty for clinical decision-making. Finally, validation across two biologically and therapeutically distinct independent cohorts demonstrated the framework’s broad clinical applicability. Ultimately, VarCP provides oncologists with mathematically reliable spatial confidence maps, marking a critical step toward safe, personalized adaptive therapy.

## Data Availability

All data produced in the present study are available upon reasonable request to the corresponding author (Stephen R. Bowen).

## CONFLICT OF INTEREST

The authors declare no conflict of interest.

## Acknowledgment

This work was supported by NIH/NCI R01CA258997.

## Notes

### Competing Interest Statement

The authors have declared no competing interest.

### Clinical Trial

NCT04151940, NCT02773238

### Author Declarations

The Institutional Review Board of the University of Washington gave ethical approval for this work, including the prospective clinical trials PET-BRIGHT (NCT04151940) and FLARE-RT (NCT02773238).

