## Supplementary file for "Uncertainty-Aware Multiscale Treatment Response Forecasting in Advanced Non-Small Cell Lung Cancer Using Residual-Variance Conformal Prediction"

**Supplementary Figure 1:** Variogram model performance for voxel level primary tumor response in FLARE RT and BRIGHT cohorts, Stable and Matérn models show the lowest weighted normalized RMSE.

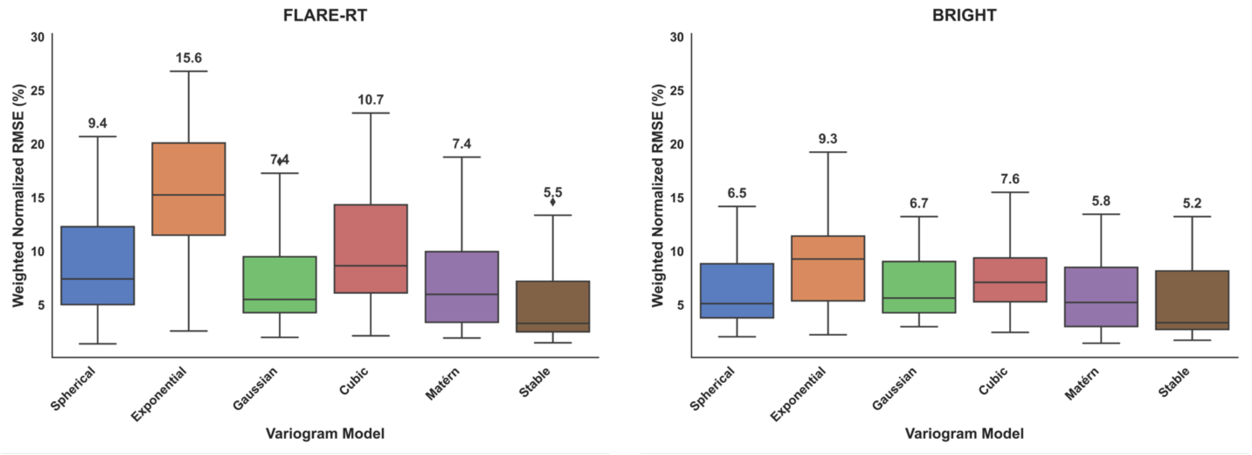


**Supplementary Table 1.** Linear Mixed-Effects Model Coefficients for Sensitivity Analysis. Each model includes a patient-level random intercept. Negative coefficients indicate lower prediction error (better performance). The selected configuration (Stable, 99% cutoff, Unweighted) maximizes spatial data utilization while maintaining a unified model across cohorts.

| | | **BRIGHT** ($n = 19$) | | **FLARE-RT** ($n = 25$) | |
|---|---|---|---|---|---|
| **Factor** | **Metric** | $\hat{\beta}$ (95% CI) | $p$ | $\hat{\beta}$ (95% CI) | $p$ |
| **Stable vs Matérn** | MAE | -0.015 (-0.016, -0.013) | $< 0.001$ | -0.005 (-0.005, -0.004) | $< 0.001$ |
| | MAE Ratio | -0.014 (-0.016, -0.012) | $< 0.001$ | -0.003 (-0.004, -0.002) | $< 0.001$ |
| | RMSE | -0.019 (-0.021, -0.018) | $< 0.001$ | -0.006 (-0.007, -0.005) | $< 0.001$ |
| | RMSE Ratio | -0.020 (-0.023, -0.017) | $< 0.001$ | -0.004 (-0.006, -0.003) | $< 0.001$ |
| **Weighted vs Unweighted** | MAE | -6.2e-06 (-1.3e-03, 1.3e-03) | 0.99 | 0.002 (0.002, 0.003) | $< 0.001$ |
| | MAE Ratio | -5.3e-05 (-1.6e-03, 1.5e-03) | 0.95 | 0.002 (0.001, 0.003) | $< 0.001$ |
| | RMSE | -1.1e-05 (-1.4e-03, 1.3e-03) | 0.99 | 0.004 (0.003, 0.005) | $< 0.001$ |
| | RMSE Ratio | -1.6e-04 (-3.1e-03, 2.7e-03) | 0.91 | 0.004 (0.002, 0.005) | $< 0.001$ |
| **Cutoff (per %)** | MAE | -2.5e-05 (-5.1e-05, 1.2e-06) | 0.06 | -2.6e-05 (-4.3e-05, -9.8e-06) | 0.002 |
| | MAE Ratio | -2.3e-05 (-5.5e-05, 9.4e-06) | 0.16 | -2.0e-05 (-3.6e-05, -3.3e-06) | 0.019 |
| | RMSE | -3.3e-05 (-6.0e-05, -5.1e-06) | 0.020 | -3.5e-05 (-5.5e-05, -1.4e-05) | $< 0.001$ |
| | RMSE Ratio | -3.3e-05 (-9.2e-05, 2.7e-05) | 0.28 | -3.0e-05 (-5.9e-05, -3.5e-07) | 0.047 |

**Supplementary Table 2.** Significant Voxel Forecast predictors (jackknife-full, p<0.05) for FLARE-RT and BRIGHT.

| **Predictor** | **FLARE-RT** | | | | **BRIGHT** | | | |
|---|---|---|---|---|---|---|---|---|
| | **Model P** | | **Model P+V** | | **Model P** | | **Model P+V** | |
| | $\beta$ (95% CI) | $p$ | $\beta$ (95% CI) | $p$ | $\beta$ (95% CI) | $p$ | $\beta$ (95% CI) | $p$ |
| $\log \mathrm{SUV}^{\mathrm{pre}}_{\mathrm{mean,lesion}}$ | 0.480 (0.226, 0.734) | $< 0.001$ | -0.379 (-0.647, -0.111) | 0.008 | 0.407 (0.120, 0.694) | 0.008 | 0.016 (-0.206, 0.237) | 0.884 |
| $\log \mathrm{SUV}^{\mathrm{pre}}_{\mathrm{voxel}}$ | — | — | 0.850 (0.738, 0.962) | $< 0.001$ | — | — | 0.426 (0.328, 0.525) | $< 0.001$ |

Jackknife-full inference: $\beta$ is the apparent (full-data) estimate; 95% CI computed as $\beta \pm t_{0.975,\, K-k_p}\, \mathrm{SE}_{\mathrm{jack}}$ from leave-one-patient-out refits.
